# Trajectories of Neurologic Recovery after Traumatic Brain Injury

**DOI:** 10.1101/2023.09.27.23296251

**Authors:** Henry E. Wang, Chengcheng Hu, Bruce J. Barnhart, Jan O. Jansen, Daniel W. Spaite

## Abstract

**BACKGROUND:** Little is known about the trajectory over time of neurologic recovery after traumatic brain injury (TBI). We sought to determine long-term changes in neurologic status after prehospital clinical trial interventions for acute TBI.

**METHODS:** We used data from the Resuscitation Outcomes Consortium Hypertonic Saline (ROC HS) TBI Trial. The trial included adult TBI, with Glasgow Coma Scale (GCS)≤8 and excluded those with shock (systolic blood pressure (SBP)≤70, or SBP 71-90 with HR≥108). The primary outcome was Glasgow Outcome Scale-Extended (GOS-E; 1=dead, 8=no disability) determined at: a) hospital discharge and b) 6-month follow-up. We analyzed changes in GOS-E between hospital discharge and 6-month follow-up by examining median changes with exact 95% confidence intervals (CI), mean changes with bootstrapped 95% CIs and Sankey graphs. We repeated the analysis for the high acuity subset of patients undergoing prehospital advanced airway insertion.

**RESULTS:** Among 1,279 TBI subjects included in the analysis, GOS-E categories at hospital discharge were: favorable (GOS-E 5-8) 220 (17.2%), unfavorable (GOS-E 2-4) 664 (51.9%), dead (GOS-E 1) 321 (25.1%), missing 74 (5.8%). GOS-E categories at 6-month follow-up were: favorable 459 (35.9%), unfavorable 279 (21.8%), dead 346 (27.1%), missing 195 (15.2%). Among initial TBI survivors with complete GOS-E, >96% followed one of three neurologic recovery trajectories: 1) favorable to favorable (20.0%), 2) unfavorable to favorable (40.3%), and 3) unfavorable to unfavorable (36.0%). Few patients deteriorated from favorable to unfavorable neurologic status and there were few additional deaths.

**CONCLUSION:** Neurologic recovery after TBI follows distinct trajectories. Among those with TBI and unfavorable neurologic status at hospital discharge, almost half will improve to favorable neurologic status at six months. Among those with favorable neurologic status at discharge, very few worsen to unfavorable neurologic status or death at six months. These findings have important implications for TBI clinical care, research and trial design.

## INTRODUCTION

The annual burden of traumatic brain injury (TBI) in the United States is enormous, associated with 2.2 million emergency department visits, 280,000 hospitalizations, 52,000 deaths, and more than $60 billion in economic costs.^1,2^ While early mitigation of secondary injury is a priority, few interventions have proven effective for TBI treatment. Clinical trials offer information to support the effectiveness of novel medical therapies. An essential consideration in clinical trial design is the selection of appropriate outcomes, which must be relevant to the disease, plausibly linked to the intervention, objectively measurable, and pertinent to patients and caregivers.

The timing of outcomes is important in TBI research.^3^ Measurement of outcomes at short-term intervals (such as hospital discharge) is feasible, but may fail to capture changes in important aspects of health status. This is highly relevant as functional recovery from TBI may take months, or even years after the initial injury. Measurement of outcomes at long-term intervals (such as 6-month follow-up) may better capture changes in health status but is logistically difficult, hence increasing trial sample size and costs. Understanding the course of health recovery is important to inform clinical trial design, but there are few studies characterizing changes in neurologic or vital status after TBI.

We sought to characterize trajectories of neurologic recovery among TBI patients enrolled in the Resuscitation Outcomes Consortium Hypertonic Saline (ROC-HS) Trial.^4^

## METHODS

### Study Design

We conducted a secondary analysis of the TBI cohort enrolled in the ROC-HS Trial. This analysis was classified as not human subjects research by the University of Arizona Institutional Review Board.

### Setting – the Resuscitation Outcomes Consortium Hypertonic Saline Trial

The ROC-HS was a multicenter clinical trial testing the effectiveness of prehospital hypertonic fluids upon outcomes after severe traumatic brain injury (TBI) and hemorrhagic shock.^4,5^ ROC consisted of 114 emergency medical services (EMS) agencies from 11 communities in the United States and Canada. The trial included enrollment of two cohorts: 1) TBI and 2) hemorrhagic shock. Inclusion criteria for the TBI cohort were: blunt mechanism of injury, age 15 years or older, Glasgow Coma Scale (GCS) score ≤8 and ineligibility for the hemorrhagic shock cohort. The hemorrhagic shock cohort included patients with systolic blood pressure of ≤70 mm Hg or 71-90 mmHg with a concomitant heart rate ≥108 beats per minute. Key exclusion criteria included known or suspected pregnancy, prisoners, transferred patients, out-of-hospital cardiopulmonary resuscitation, administration of >2000 ml of crystalloid intravenous fluid or any amount of colloid or blood products. Patients with concomitant head injury and hypotension were included in the shock cohort.

The three blinded trial interventions included a 250 ml bolus of: 1) 7.5% saline (hypertonic saline), 2) 7.5% saline with 6% dextran 70 (hypertonic saline/dextran), and 3) 0.9% saline (normal saline), randomized in a ratio of 1:1:1.4. Study fluids were provided in identical intravenous bags; EMS providers were blinded to the fluid contents. The trial met prespecified criteria for futility after enrollment of 1,282 TBI and 853 hemorrhagic shock. Patient enrollment occurred during 2006-2009.

### Selection of Participants

We included all patients enrolled in the TBI cohort of the parent trial.

### Outcomes

The outcomes of interest for this analysis were neurologic outcome and death at hospital discharge and 6-month follow-up. The trial determined neurologic status using the Glasgow Outcome Score-Extended (GOSE).^6,7^ The Glasgow Outcome Scale (GOS) was first described by Jennett and Bond as an assessment of global neurologic outcome and death after severe brain injury.^8^ Later extensions improved its sensitivity.^6,7^ Research teams determined GOS-E at hospital discharge using a structured telephone survey.^9^ If the patient was unable to respond to the survey, family members or caregivers provided the requested information, an approach that has been previously validated.^9,10^ If the neurologic status survey occurred after six months, we considered the assessment to have occurred at six months. For patients who died after hospital discharge, we assigned a 6-month GOS-E=1. For patients who died before hospital discharge, we assigned GOS-E=1 for both hospital and 6-month follow-up.

### Data Analysis

We excluded patients where hospital discharge occurred after the 6-month follow-up. We assessed characteristics of the study population, including patient demographics (age, sex), clinical presentation (injury mechanism, prehospital Glasgow Coma Scale, revised trauma score (RTS), injury severity score (ISS), head abbreviated injury score (AIS)), EMS care (air medical transport, advanced airway placement, the prehospital time interval), and outcomes (hospital survival, 28-day survival, intensive care unit-free days at 28 days, and GOS-E at hospital discharge and six months). We determined GOS-E changes between hospital discharge and six months by examining median changes with exact 95% CIs and means change with bootstrapped 95% CIs. We stratified neurologic status according to severity categories: GOS-E 1 = dead, 2-4 = unfavorable neurologic status, and 5-8 = favorable neurologic status. We also used Sankey graphs to depict transitions between GOS-E categories at hospital discharge and six months.

We repeated the analysis limited to the higher acuity subset of patients undergoing prehospital advanced airway management (endotracheal intubation, supraglottic airways or surgical airways). In an additional analysis, we focused on the subset of patients with unfavorable neurologic status at hospital discharge, distinguishing those who improved to favorable neurologic status with those who remained in unfavorable neurologic status at 6-month follow-up. Using multi-covariate logistic regression, we identified the baseline characteristics independently associated with improvement to good 6-month neurologic function through a backward elimination process starting from a model including all baseline characteristics that were associated with the outcome in univariate analysis at the significance level of 0.20.

Some authors advocate extending the GOS-E range to 4-8 for classifying favorable neurologic status, noting that many patients with GOS-E=4 (upper severe disability) are able to function at home without supervision for more than 8 hours daily.^11^ In a sensitivity analysis, we repeated the analysis of the full cohort defining favorable neurologic status as GOS-E 4-8 and unfavorable neurologic status as GOS-E 2-3.

We conducted all analyses using SAS (Cary, North Carolina) and Microsoft Excel (Redmond, Washington) with the Power-User add-in (Power-User SAS, Paris, France).

## RESULTS

The parent trial enrolled a total of 1,282 TBI patients. We excluded three patients who were discharged from the hospital more than six months after injury, leaving 1,279 in the analysis. Trial interventions were: hypertonic saline+dextrose 357 (27.9%), hypertonic saline 340 (26.6%), and normal saline 582 (45.5%). Enrolled subjects were mostly male and suffered primarily blunt injury (Table 1). The acuity of the population was high, with high RTS and ISS. Approximately 60% received successful prehospital advanced airway insertion, and approximately 40% underwent air medical transport. One-third of the population experienced severe TBI (head AIS 3 or 4), and one-third suffered critical TBI (head AIS 5-6); one-fifth of the population did not have a significant brain injury. Approximately three-fourths were alive at 28-days after injury. Among the 945 patients alive at hospital discharge, median time from injury to discharge was 14 days (IQR 4, 30), and median time from hospital discharge to 6-month outcome interview was 176 days (IQR 158, 190).

**TABLE 1.**
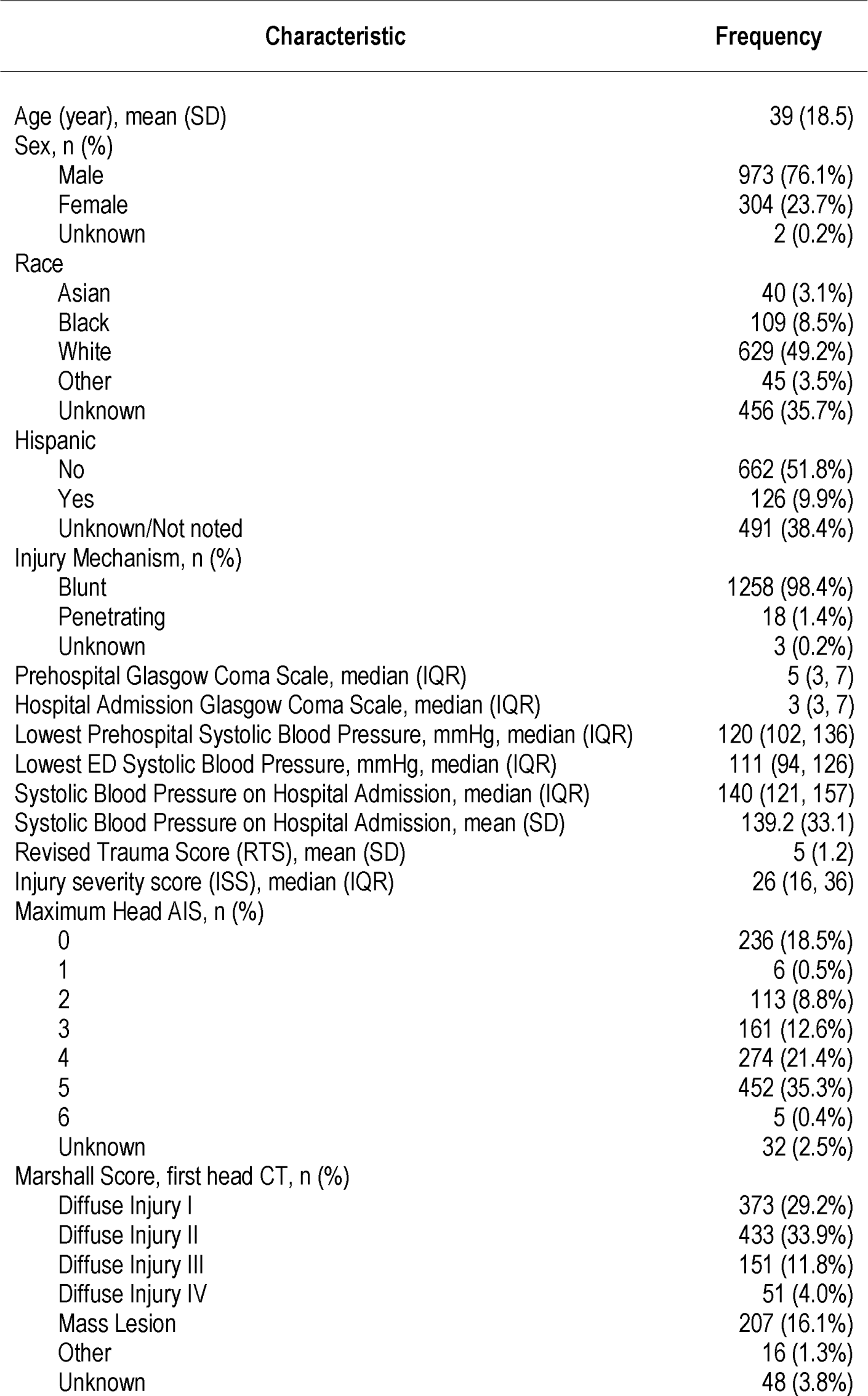
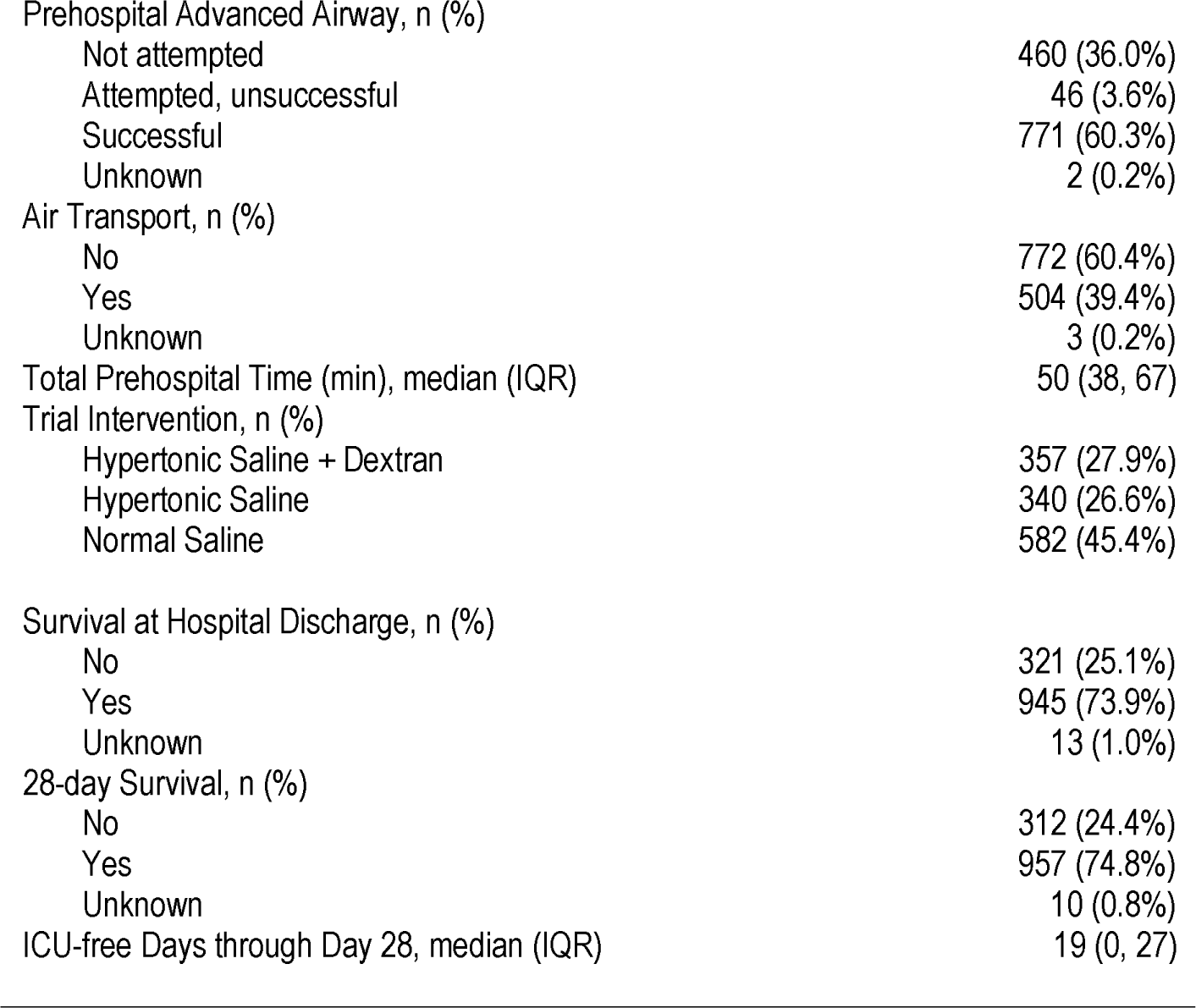
Characteristics of 1,279 patients enrolled in the Resuscitation Outcomes Consortium Hypertonic Saline Traumatic Brain Injury (TBI) trial.

| Characteristic | Frequency |
| --- | --- |
| Age (year), mean (SD) | 39 (18.5) |
| Sex, n (%) |  |
| Male | 973 (76.1%) |
| Female | 304 (23.7%) |
| Unknown | 2 (0.2%) |
| Race |  |
| Asian | 40 (3.1%) |
| Black | 109 (8.5%) |
| White | 629 (49.2%) |
| Other | 45 (3.5%) |
| Unknown | 456 (35.7%) |
| Hispanic |  |
| No | 662 (51.8%) |
| Yes | 126 (9.9%) |
| Unknown/Not noted | 491 (38.4%) |
| Injury Mechanism, n (%) |  |
| Blunt | 1258 (98.4%) |
| Penetrating | 18 (1.4%) |
| Unknown | 3 (0.2%) |
| Prehospital Glasgow Coma Scale, median (IQR) | 5 (3, 7) |
| Hospital Admission Glasgow Coma Scale, median (IQR) | 3 (3, 7) |
| Lowest Prehospital Systolic Blood Pressure, mmHg, median (IQR) | 120 (102, 136) |
| Lowest ED Systolic Blood Pressure, mmHg, median (IQR) | 111 (94, 126) |
| Systolic Blood Pressure on Hospital Admission, median (IQR) | 140 (121, 157) |
| Systolic Blood Pressure on Hospital Admission, mean (SD) | 139.2 (33.1) |
| Revised Trauma Score (RTS), mean (SD) | 5 (1.2) |
| Injury severity score (ISS), median (IQR) | 26 (16, 36) |
| Maximum Head AIS, n (%) |  |
| 0 | 236 (18.5%) |
| 1 | 6 (0.5%) |
| 2 | 113 (8.8%) |
| 3 | 161 (12.6%) |
| 4 | 274 (21.4%) |
| 5 | 452 (35.3%) |
| 6 | 5 (0.4%) |
| Unknown | 32 (2.5%) |
| Marshall Score, first head CT, n (%) |  |
| Diffuse Injury I | 373 (29.2%) |
| Diffuse Injury II | 433 (33.9%) |
| Diffuse Injury III | 151 (11.8%) |
| Diffuse Injury IV | 51 (4.0%) |
| Mass Lesion | 207 (16.1%) |
| Other | 16 (1.3%) |
| Unknown | 48 (3.8%) |
| Prehospital Advanced Airway, n (%) |  |
| Not attempted | 460 (36.0%) |
| Attempted, unsuccessful | 46 (3.6%) |
| Successful | 771 (60.3%) |
| Unknown | 2 (0.2%) |
| Air Transport, n (%) |  |
| No | 772 (60.4%) |
| Yes | 504 (39.4%) |
| Unknown | 3 (0.2%) |
| Total Prehospital Time (min), median (IQR) | 50 (38, 67) |
| Trial Intervention, n (%) |  |
| Hypertonic Saline + Dextran | 357 (27.9%) |
| Hypertonic Saline | 340 (26.6%) |
| Normal Saline | 582 (45.4%) |
| Survival at Hospital Discharge, n (%) |  |
| No | 321 (25.1%) |
| Yes | 945 (73.9%) |
| Unknown | 13 (1.0%) |
| 28-day Survival, n (%) |  |
| No | 312 (24.4%) |
| Yes | 957 (74.8%) |
| Unknown | 10 (0.8%) |
| ICU-free Days through Day 28, median (IQR) | 19 (0, 27) |

At hospital discharge, favorable neurologic status, unfavorable neurologic status and death comprised 17.2%, 51.9% and 25.1% of the full cohort. (Table 2) At 6-month follow-up favorable neurologic status, unfavorable neurologic status and death comprised 35.9%, 21.8% and 27.1% of the cohort.

**TABLE 2.** Comparison of neurologic status (Glasgow Outcome Scale-Extended - GOS-E) at hospital discharge and 6-month follow-up among patients enrolled in the Resuscitation Outcomes Consortium Hypertonic Saline Traumatic Brain Injury (TBI) trial. Includes full cohort (n=1,279). Deaths assigned GOS-E=1. GOS-E 2-4 classified as unfavorable neurologic status. GOS-E 5-8 classified as favorable neurologic status. Shaded cells indicate transitions between favorable and unfavorable neurologic status or death.

|  |  | GOS-E Classification<br>GOS-E at 6-Month Follow-Up |  |  |  |  |  |  |  |  | TOTAL |
| --- | --- | --- | --- | --- | --- | --- | --- | --- | --- | --- | --- |
|  |  | Favorable<br>n=459 (35.9%) |  |  |  | Unfavorable<br>n=279 (21.8%) |  |  | Dead<br>n=346<br>(27.1%) | Missing<br>n=195<br>(15.2%) |  |
| GOS-E Classification | GOS-E at Hospital Discharge | 8 | 7 | 6 | 5 | 4 | 3 | 2 | 1 | Missing |  |
| Favorable<br>n=200<br>(17.2%) | 8 | 57 | 9 | 6 | 3 | 2 | 0 | 0 | 0 | 39 | 116 |
|  | 7 | 17 | 4 | 1 | 3 | 0 | 0 | 0 | 0 | 9 | 34 |
|  | 6 | 20 | 6 | 3 | 6 | 1 | 0 | 0 | 1 | 12 | 49 |
|  | 5 | 4 | 7 | 2 | 1 | 1 | 1 | 0 | 1 | 4 | 21 |
| Unfavorable<br>n=664<br>(51.9%) | 4 | 35 | 16 | 9 | 11 | 17 | 8 | 0 | 0 | 14 | 110 |
|  | 3 | 78 | 48 | 58 | 44 | 77 | 131 | 1 | 9 | 52 | 498 |
|  | 2 | 1 | 0 | 0 | 0 | 3 | 25 | 6 | 12 | 9 | 56 |
| Dead n=321<br>(25.1%) | 1 | 0 | 0 | 0 | 0 | 0 | 0 | 0 | 321 | 0 | 321 |
| Missing n=74<br>(5.8%) | Missing | 6 | 2 | 1 | 1 | 1 | 4 | 1 | 2 | 56 | 74 |
| TOTAL |  | 218 | 92 | 80 | 69 | 102 | 169 | 8 | 346 | 195 | 1,279 |

No change between favorable/unfavorable status or death Improvement in neurologic status Decline in neurologic status or death

Neurologic status was available at both hospital discharge and 6-month follow-up for 1,066 of 1,279 (83.3%) subjects. Of this subset, 30.1% were dead at hospital discharge. Of the remaining TBI survivors most (>96%) followed one of three neurologic status trajectories: 1) favorable to favorable (20.0%), 2) unfavorable to favorable (40.3%), and 3) unfavorable to unfavorable (36.0%). (Table 3, Figure 1). Of the 220 patients with favorable neurologic status at hospital discharge, only 7 (1.0%) deteriorated to unfavorable neurologic status or death. Compared with hospital discharge, there were only 23 additional deaths at 6-month follow-up.

**FIGURE 1.**
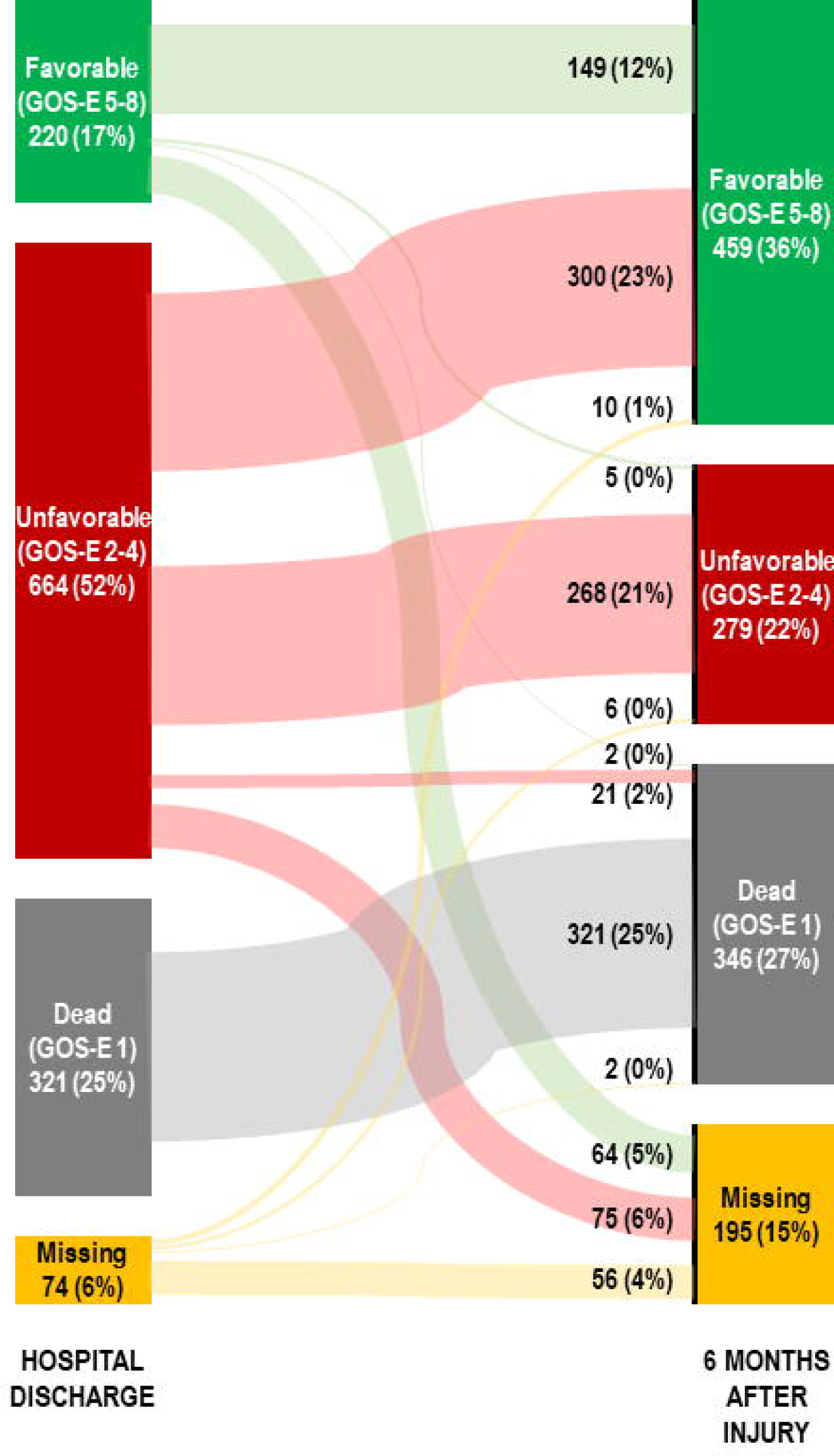
Sankey graph depicting changes in neurologic status (Glasgow Outcome Scale-Extended - GOS-E) between hospital discharge and 6-month follow-up among patients enrolled in the Resuscitation Outcomes Consortium Hypertonic Saline Trial. N=1,279 patients.

**TABLE 3.** Trajectories of neurologic status between hospital discharge and 6-month follow-up among patients enrolled in the Resuscitation Outcomes Consortium Hypertonic Saline Traumatic Brain Injury (TBI) trial. Includes 1,066 of 1,279 TBI subjects with complete neurologic status at both hospital discharge and 6-month follow-up. Neurologic status defined as: favorable GOS-E 5-8, unfavorable GOS-E 2-4, dead GOS-E 1.

| Neurologic Trajectory<br>(Hospital Discharge → 6-Month Follow-Up) | Full Cohort<br>N (%) | Hospital<br>Survivors Only<br>N (%) |
| --- | --- | --- |
| 1) Favorable (GOS-E 5-8) → Favorable (GOS-E 5-8) | 149 (14.0%) | 149 (20.0%) |
| 2) Favorable (GOS-E 5-8) → Unfavorable (GOS-E 2-4) | 5 (0.5%) | 5 (0.7%) |
| 3) Favorable (GOS-E 5-8) → Dead (GOS-E 1) | 2 (0.2%) | 2 (0.3%) |
| 4) Unfavorable (GOS-E 2-4) → Favorable (GOS-E 5-8) | 300 (28.1%) | 300 (40.3%) |
| 5) Unfavorable (GOS-E 2-4) → Unfavorable (GOS-E 2-4) | 268 (25.1%) | 268 (36.0%) |
| 6) Unfavorable (GOS-E 2-4) → Dead (GOS-E 1) | 21 (2.0%) | 21 (2.8%) |
| 7) Dead at Hospital Discharge (GOS-E 1) | 321 (30.1%) | N/A |

At hospital discharge, favorable neurologic status, unfavorable neurologic status and death comprised 12.5%, 56.2% and 28.5% of the advanced airway cohort. (Appendix 1) At 6-month follow-up favorable neurologic status, unfavorable neurologic status or death comprised 35.9%, 22.7% and 30.1% of the cohort.

Neurologic status was available at both hospital discharge and 6-month follow-up for 679 of 771 (88.0%) advanced airway subjects. Of this subset, 32.4 % were dead at hospital discharge. Of the remaining TBI survivors, most (>97%) followed one of three neurologic status trajectories: 1) favorable to favorable (14.4%), 2) unfavorable to favorable (45.3%), and 3) unfavorable to unfavorable (37.7%). (Appendices 2 and 3). Of the 220 patients with favorable neurologic status at hospital discharge, only 2 (0.4%) deteriorated to unfavorable neurologic status or death. Compared with hospital discharge, there were only 12 additional deaths at 6-month follow-up.

In the full cohort, younger age, higher Revised Trauma Score, lower head AIS score, lower Marshall Score, and absence of hypotension at ED were independently associated with progression from unfavorable neurologic status (GOS-E 2-4) at hospital discharge to good neurologic status (GOS-E 5-8) at 6-month follow-up. (Appendix 4, Table 4)

**TABLE 4.** Multi-covariate logistic regression of factors associated with improvement from unfavorable (GOS-E 2-4) to favorable neurologic outcome (GOS-E 5-8) among patients enrolled in the Resuscitation Outcomes Consortium Hypertonic Saline Traumatic Brain Injury (TBI) trial. Limited to subset with unfavorable neurologic status at hospital discharge (GOS-E 2-4).

| Variable | OR (95% CI) |
| --- | --- |
| **Age (year)** | 0.97 (0.96, 0.98) |
| **Revised Trauma Score (RTS)** | 1.44 (1.20, 1.72) |
| **Maximum Head AIS** |  |
| 0 | Reference |
| 1-3 | 1.40 (0.75, 2.62) |
| 4 | 1.06 (0.57, 1.97) |
| 5 | 0.61 (0.32, 1.14) |
| **Marshall Score, first head CT** |  |
| Diffuse Injury I/II | Reference |
| Diffuse Injury III/IV | 0.50 (0.29, 0.85) |
| Mass Lesion | 0.52 (0.28, 0.98) |
| **Hypotension at ED (lowest SBP < 90 mmHg)** |  |
| No | Reference |
| Yes | 0.56 (0.34, 0.94) |

In a sensitivity analysis recategorizing favorable neurologic status as GOS-E 4-8 and unfavorable neurologic status as GOS-E 2-3, there were only minimal differences in the percentage of surviving patients transitioning from unfavorable to favorable neurologic status (41.5%). (Appendices 5 and 6)

## DISCUSSION

In this analysis of data from the ROC-HS trial, the majority of TBI patients alive at hospital discharge subsequently exhibited one of three neurologic recovery trajectories: 1) good neurologic status at both hospital discharge and 6-month follow-up, 2) poor neurologic status at hospital discharge improving to good neurologic outcome at 6-month follow-up, and 3) poor neurologic status at both hospital discharge and 6-month follow-up. Among those with initially unfavorable neurologic status, almost half improved to favorable neurologic status. Very few patients with favorable neurologic status at hospital discharge deteriorated or died at 6-month follow-up. There were also few additional deaths at 6-month follow-up.

Our findings have important implications for TBI research and clinical trial design. If the objective of a novel intervention is to mitigate death after TBI, then hospital survival may be suitable as the primary research outcome, as the overall number of deaths does not significantly increase at six months. For interventions targeting optimization of neurologic outcome, extending observation to six months or later may be necessary. However, researchers can likely focus attention on the subset with unfavorable neurologic status at hospital discharge, as half of this group is likely to improve to favorable neurologic status at six months. It also appears that if differences in neurologic outcome are observed at hospital discharge, one may expect even larger widening of effect at later time points. Similarly, our observations suggest that studies focused on TBI rehabilitation will likely target only half of the total TBI population; - i.e., those with unfavorable neurologic status at hospital discharge.

Our findings also reinforce existing perspectives regarding the trajectories of TBI neurologic recovery. As suggested by the current and prior studies, the general ominous prognosis of patients with poor neurologic status at hospital discharge may be unwarranted; over the 6-24 months following TBI, almost half of patients will improve from unfavorable to favorable neurologic status.^11–16^ For example, in an analysis of 484 patients from the TRACK-TBI cohort study, McCrea, et al. found that at 12 months, 52% of patients with severe TBI (defined as initial GCS 3-8) achieved favorable neurologic outcomes.^11^ The distinction of our study is that the data specifically originated from TBI patients enrolled in a clinical trial. Among initial survivors at hospital discharge, we observed only 23 additional deaths at six months, representing 1.8% of the study population, rates much lower than in prior cohorts.^11,13^ This may be due to the fact that a TBI clinical trial population will invariably include select patients without head CT abnormalities. A strength of the current analysis is that the rates of GOS-E missingness were 6-15%, much lower than the 30-50% missingness observed in prior cohorts.^11–13^ These observations highlight the importance of strategies to mitigate and manage missing outcomes in TBI trials.

We also offer a novel analytic approach to better depict and conceptualize neurologic recovery after TBI. Prior studies presented the distribution of GOS-E categories at discrete time points, characterizing the heterogeneity of the population but not specifically identifying the transitions of neurological status between discharge and follow-up.^11–13^ Our use of Sankey graphs addresses this limitation by depicting both the distributions of, and the transitions between, GOS-E categories. Other studies have characterized TBI trajectories using a range of sophisticated analytic techniques, such as quadratic models and least absolute shrinkage and selection operator (LASSO).^14,16^ Our approach is simpler, more intuitive, and clarifies that neurologic recovery after TBI entails seven potential trajectories, of which four are dominant (Table 3). Among these trajectories, the transition from unfavorable to favorable neurologic status was the largest cohort and of greatest interest. We believe these results will be helpful for designing TBI studies and trials, focusing attention and resources on the subset of patients with initial unfavorable neurologic status. This may provide an efficient approach since it appears that both death and neurological deterioration are very infrequent after discharge.

## LIMITATIONS

Our observations must be interpreted in light of the design of the parent trial. The ROC data are now almost 15 years old, and repeating this analysis in a more recent data set is merited. The ROC-HS Trial included high acuity TBI with an initial presenting GCS ≤8. Thus, the trial may have included some unconscious subjects without intracranial injuries (for example, those with intoxication masquerading as TBI) as well as seemingly conscious patients with severe intracranial injuries. Missingness of GOS-E was also prevalent, influencing 17% of the cohort. If all the missing values were included in sensitivity analyses as “favorable” or “unfavorable” neurologic status, this would have slightly shifted the proportion of patients in each neurologic trajectory. We did not study other measures of neurologic and functional status such as the Functional Independence Measure, the Disability Rating Scale or quality of life.^3,17^ We also did not study other neurologic sequelae such as seizures, dementia and Parkinsonism.^18^

## CONCLUSION

Neurologic recovery after TBI follows distinct trajectories. Among TBI patients with unfavorable neurologic status at hospital discharge, almost half will progress to favorable neurologic status at six months. Very few TBI victims experience worsened neurologic status or death at six months. These observations offer key perspectives to guide TBI research and clinical trial design.

## TRANSPARENCY, RIGOR AND REPRODUCIBILITY

This study was a secondary analysis of existing data resulting from the Resuscitation Outcomes Consortium Hypertonic Saline Traumatic Brain Injury Trial. The parent trial was registered at clinicaltrials.gov (NCT00316004). The analysis plan for this secondary analysis was not formally pre-registered. Sample size estimates were not determined for this secondary analysis. The parent trial enrolled a total of 1,282 TBI patients, of which 1,279 with complete outcomes were included in this secondary analysis. The analysis was carried out on a de-identified data set without linkage to individual subjects. Data were acquired from the NHLBI BioLINCC repository. The software used to perform the analyses (SAS and Microsoft Excel) are widely available. The key inclusion criteria (e.g., primary diagnosis or prognostic factor) are established standards in the field. The sample sizes and degrees of freedom reflect the number of independent measurements (i.e., number of individual participants). No replication or external validation studies have been performed or are planned/ongoing at this time to our knowledge. Data for this study are available from the NHLBI BioLINCC. This article will not be available for Open Access.

## Data Availability

Data for this study may be requested from the NHLBI BioLINCC repository.

## ACKNOWLEDGEMENTS

This analysis was prepared using research materials obtained from the National Heart Lung and Blood Institute (NHLBI) Biologic Specimen and Data Repository Information Coordinating Center (BioLINCC) and does not necessarily reflect the opinions or views of NHLBI.

## PRESENTATION

Presented at the National Associated of EMS Physicians Annual Meeting, Tampa, Florida, January 27, 2023.

## AUTHOR CONTRIBUTIONS

All authors have made substantial contributions to all of the following: (1) the conception and design of the study, or acquisition of data, or analysis and interpretation of data, (2) drafting the article or revising it critically for important intellectual content, (3) final approval of the version to be submitted.

## REFERENCES

1. Finkelstein E, Corso PS, Miller TR. The incidence and economic burden of injuries in the United States. Oxford; New York: Oxford University Press; 2006.

2. Corrigan JD, Selassie AW, Orman JA. The epidemiology of traumatic brain injury. J Head Trauma Rehabil. 2010;25(2):72–80.

3. Shukla D, Devi BI, Agrawal A. Outcome measures for traumatic brain injury. Clinical Neurology and Neurosurgery. 2011;113(6):435–441.

4. Bulger EM, May S, Brasel KJ, et al. Out-of-hospital hypertonic resuscitation following severe traumatic brain injury: a randomized controlled trial. JAMA. 2010;304(13):1455–1464.

5. Bulger EM, May S, Kerby JD, et al. Out-of-hospital hypertonic resuscitation after traumatic hypovolemic shock: a randomized, placebo controlled trial. Annals of surgery. 2011;253(3):431–441.

6. Wilson JT, Pettigrew LE, Teasdale GM. Structured interviews for the Glasgow Outcome Scale and the extended Glasgow Outcome Scale: guidelines for their use. J Neurotrauma. 1998;15(8):573–585.

7. Jennett B, Snoek J, Bond MR, Brooks N. Disability after severe head injury: observations on the use of the Glasgow Outcome Scale. J Neurol Neurosurg Psychiatry. 1981;44(4):285–293.

8. Jennett B, Bond M. Assessment of outcome after severe brain damage. Lancet. 1975;1(7905):480-484.

9. Pettigrew LE, Wilson JT, Teasdale GM. Assessing disability after head injury: improved use of the Glasgow Outcome Scale. J Neurosurg. 1998;89(6):939–943.

10. Hall K, Cope DN, Rappaport M. Glasgow Outcome Scale and Disability Rating Scale: comparative usefulness in following recovery in traumatic head injury. Arch Phys Med Rehabil. 1985;66(1):35–37.

11. McCrea MA, Giacino JT, Barber J, et al. Functional Outcomes Over the First Year After Moderate to Severe Traumatic Brain Injury in the Prospective, Longitudinal TRACK-TBI Study. JAMA Neurol. 2021;78(8):982–992.

12. Puffer RC, Yue JK, Mesley M, et al. Recovery Trajectories and Long-Term Outcomes in Traumatic Brain Injury: A Secondary Analysis of the Phase 3 Citicoline Brain Injury Treatment Clinical Trial. World Neurosurg. 2019;125:e909–e915.

13. Wilkins TE, Beers SR, Borrasso AJ, et al. Favorable Functional Recovery in Severe Traumatic Brain Injury Survivors beyond Six Months. J Neurotrauma. 2019;36(22):3158–3163.

14. Pretz CR, Dams-O’Connor K. Longitudinal description of the glasgow outcome scale-extended for individuals in the traumatic brain injury model systems national database: a National Institute on Disability and Rehabilitation Research traumatic brain injury model systems study. Arch Phys Med Rehabil. 2013;94(12):2486–2493.

15. Puffer RC, Yue JK, Mesley M, et al. Long-term outcome in traumatic brain injury patients with midline shift: a secondary analysis of the Phase 3 COBRIT clinical trial. J Neurosurg. 2018;131(2):596–603.

16. Rubin ML, Yamal JM, Chan W, Robertson CS. Prognosis of Six-Month Glasgow Outcome Scale in Severe Traumatic Brain Injury Using Hospital Admission Characteristics, Injury Severity Characteristics, and Physiological Monitoring during the First Day Post-Injury. J Neurotrauma. 2019;36(16):2417–2422.

17. Linacre JM, Heinemann AW, Wright BD, Granger CV, Hamilton BB. The structure and stability of the functional independence measure. Archives of Physical Medicine and Rehabilitation. 1994;75(2):127–132.

18. Bazarian JJ, Cernak I, Noble-Haeusslein L, Potolicchio S, Temkin N. Long-term neurologic outcomes after traumatic brain injury. J Head Trauma Rehabil. 2009;24(6):439–451.

19. Maiden MJ, Cameron PA, Rosenfeld JV, Cooper DJ, McLellan S, Gabbe BJ. Long-Term Outcomes after Severe Traumatic Brain Injury in Older Adults. A Registry-based Cohort Study. Am J Respir Crit Care Med. 2020;201(2):167–177.19.

