## Supplemental Files for "Trajectories of Neurologic Recovery after Traumatic Brain Injury"

Supplemental Appendices

July 9, 2023

### APPENDIX 1

Comparison of neurologic status (Glasgow Outcome Scale-Extended - GOS-E) at hospital discharge and 6-month follow-up among patients enrolled in the Resuscitation Outcomes Consortium Hypertonic Saline Traumatic Brain Injury (TBI) trial. Prehospital advanced airway cohort (n=771). Deaths assigned GOS-E=1. GOS-E 2-4 classified as unfavorable neurologic status. GOS-E 5-8 classified as favorable neurologic status. Shaded cells indicate transitions between favorable and unfavorable neurologic status or death.

|  |  | GOS-E Classification  GOS-E at 6-Month Follow-Up | | | | | | | | |  |
| --- | --- | --- | --- | --- | --- | --- | --- | --- | --- | --- | --- |
|  |  | **Favorable**  n=277 (35.9%) | | | | **Unfavorable**  n=175 (22.7%) | | | Dead n=232 (30.1%) | Missing n=87 (11.3%) |  |
| GOS-E Classification | GOS-E at Hospital Discharge | 8 | 7 | 6 | 5 | 4 | 3 | 2 | 1 | Missing | TOTAL |
| **Favorable**  n=96 (12.5%) | 8 | 24 | 4 | 2 | 1 | 0 | 0 | 0 | 0 | 16 | **47** |
|  | 7 | 7 | 3 | 1 | 1 | 0 | 0 | 0 | 0 | 4 | **16** |
|  | 6 | 10 | 3 | 1 | 2 | 1 | 0 | 0 | 1 | 7 | **25** |
|  | 5 | 3 | 4 | 0 | 0 | 0 | 0 | 0 | 0 | 1 | **8** |
| **Unfavorable**  n=433 (51.9%) | 4 | 24 | 11 | 6 | 5 | 7 | 4 | 0 | 0 | 7 | **64** |
|  | 3 | 47 | 36 | 47 | 31 | 53 | 79 | 1 | 2 | 28 | **324** |
|  | 2 | 1 | 0 | 0 | 0 | 3 | 21 | 5 | 8 | 7 | **45** |
| Dead n=220 (28.5%) | 1 | 0 | 0 | 0 | 0 | 0 | 0 | 0 | 220 | 0 | **220** |
| Missing n=22 (2.9%) | Missing | 2 | 1 | 0 | 0 | 0 | 1 | 0 | 1 | 17 | **22** |
|  | TOTAL | **118** | **62** | **57** | **40** | **64** | **105** | **6** | **232** | **87** | **771** |

|  | No change between favorable/unfavorable status or death |
| --- | --- |
|  | Improvement in neurologic status |
|  | Decline in neurologic status or death |

### APPENDIX 2

Sankey graph depicting changes in neurologic status (GOS-E) between hospital discharge and 6-month follow-up among patients enrolled in the Resuscitation Outcomes Consortium Hypertonic Saline Traumatic Brain Injury (TBI) trial. Limited to patients receiving prehospital advanced airway management, n=771.

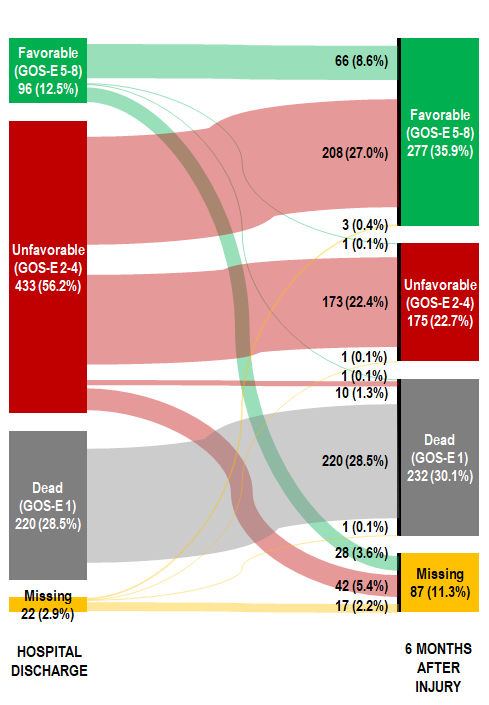

### APPENDIX 3

Trajectories of neurologic status between hospital discharge to 6-month follow-up among patients enrolled in the Resuscitation Outcomes Consortium Hypertonic Saline Traumatic Brain Injury (TBI) trial. Includes only the 679 of 771 TBI receiving prehospital advanced airway management with complete neurologic status at both hospital discharge and 6-month follow-up.

| **Neurologic Status Trajectory**  **(Hospital Discharge 🡪 6-Month Follow-Up)** | **Advanced Airway Cohort**  **N (%)** | **Advanced Airway Cohort, Hospital Survivors Only**  **N (%)** |
| --- | --- | --- |
| 1) Favorable 🡪 Favorable | 66 (9.7%) | 66 (14.4%) |
| 2) Favorable 🡪 Unfavorable | 1 (0.1%) | 1 (0.2%) |
| 3) Favorable 🡪 Dead | 1 (0.1%) | 1 (0.2%) |
| 4) Unfavorable 🡪 Favorable | 208 (30.6%) | 208 (45.3%) |
| 5) Unfavorable 🡪 Unfavorable | 173 (25.5%) | 173 (37.7%) |
| 6) Unfavorable 🡪 Dead | 10 (1.5%) | 10 (2.2%) |
| 7) Dead at Hospital Discharge | 220 (32.4%) | N/A |

### APPENDIX 4

Characteristics of patients discharged from hospital with unfavorable neurologic status, stratified by change in 6-month neurologic status. Include patients enrolled in the Resuscitation Outcomes Consortium Hypertonic Saline Traumatic Brain Injury (TBI) trial. Unfavorable neurologic status=Glasgow Outcome Scale Extended 2-4. Favorable neurologic status=Glasgow Outcome Scale Extended 5-8.

| **Characteristic** | **Unfavorable 🡪 Unfavorable**  **N=289** | **Unfavorable 🡪 Favorable**  **N=300** | **p-value** |
| --- | --- | --- | --- |
| Age (year), mean (SD) | 39.7 (18) | 34 (15.2) | <0.001 |
| Sex, n (%) |  |  | 0.87 |
| Male | 216 (74.7%) | 227 (75.7%) |  |
| Female | 73 (25.3%) | 73 (24.3%) |  |
| Race |  |  | 0.74 |
| Asian | 13 (4.5%) | 10 (3.3%) |  |
| Black | 24 (8.3%) | 27 (9%) |  |
| White | 150 (51.9%) | 170 (56.7%) |  |
| Other | 11 (3.8%) | 9 (3%) |  |
| Unknown | 91 (31.5%) | 84 (28%) |  |
| Hispanic |  |  | 0.22 |
| No | 165 (57.1%) | 172 (57.3%) |  |
| Yes | 21 (7.3%) | 33 (11%) |  |
| Unknown/Not noted | 103 (35.6%) | 95 (31.7%) |  |
| Injury Mechanism, n (%) |  |  | 0.36 |
| Blunt | 286 (99%) | 299 (99.7%) |  |
| Penetrating | 3 (1%) | 1 (0.3%) |  |
| Prehospital Glasgow Coma Scale, median (IQR) | 5 (3, 7) | 6 (3, 7) | 0.01 |
| Hospital Admission Glasgow Coma Scale, median (IQR) | 3 (3, 6) | 3 (3, 7) | 0.86 |
| Lowest Prehospital Systolic Blood Pressure, mmHg, median (IQR) | 116.5 (100, 134) | 120 (107, 135) | 0.11 |
| Lowest ED Systolic Blood Pressure, mmHg, median (IQR) | 110 (94, 125) | 113 (100, 127) | 0.04 |
| Systolic Blood Pressure on Hospital Admission, median (IQR) | 140 (121, 160) | 142 (127.5, 157) | 0.18 |
| Revised Trauma Score (RTS), mean (SD) | 4.9 (1) | 5.3 (1.1) | < 0.001 |
| Injury severity score (ISS), median (IQR) | 29 (22, 38) | 25.5 (17, 34) | < 0.001 |
| Maximum Head AIS, n (%) |  |  | < 0.001 |
| 0 | 36 (12.5%) | 41 (13.7%) |  |
| 1 | 0 (0%) | 1 (0.3%) |  |
| 2 | 12 (4.2%) | 30 (10%) |  |
| 3 | 39 (13.5%) | 61 (20.3%) |  |
| 4 | 80 (27.7%) | 91 (30.3%) |  |
| 5 | 116 (40.1%) | 73 (24.3%) |  |
| 6 | 0 (0%) | 0 (0%) |  |
| Unknown | 6 (2.1%) | 3 (1%) |  |
| Marshall Score, first head CT, n (%) |  |  | < 0.001 |
| Diffuse Injury I | 53 (18.3%) | 92 (30.7%) |  |
| Diffuse Injury II | 140 (48.4%) | 146 (48.7%) |  |
| Diffuse Injury III | 42 (14.5%) | 33 (11%) |  |
| Diffuse Injury IV | 12 (4.2%) | 4 (1.3%) |  |
| Mass Lesion | 39 (13.5%) | 22 (7.3%) |  |
| Other | 3 (1%) | 1 (0.3%) |  |
| Unknown | 0 (0%) | 2 (0.7%) |  |
| Prehospital Advanced Airway, n (%) |  |  | 0.23 |
| Not attempted | 96 (33.2%) | 86 (28.7%) |  |
| Attempted, unsuccessful | 10 (3.5%) | 6 (2%) |  |
| Successful | 183 (63.3%) | 208 (69.3%) |  |
| Air Transport, n (%) |  |  | 0.87 |
| No | 156 (54%) | 165 (55%) |  |
| Yes | 133 (46%) | 135 (45%) |  |
| Total Prehospital Time (min), median (IQR) | 52.2 (39, 69.9) | 53 (39.5, 71.6) | 0.67 |
| Trial Intervention, n (%) |  |  | 0.59 |
| Hypertonic Saline + Dextran | 86 (29.8%) | 81 (27%) |  |
| Hypertonic Saline | 81 (28%) | 80 (26.7%) |  |
| Normal Saline | 122 (42.2%) | 139 (46.3%) |  |
| Fisher’s exact test or Chi-squared test for un-ordered categorical variables and Kruskal-Wallis test for continuous or ordinal categorical variables; the unknown category, if present, is excluded from the testing procedure. | | | |

### APPENDIX 5

Sensitivity analysis. Comparison of neurologic status (Glasgow Outcome Scale-Extended - GOS-E) at hospital discharge and 6-month follow-up among patients enrolled in the Resuscitation Outcomes Consortium Hypertonic Saline Traumatic Brain Injury (TBI) trial. Include

full cohort (n=1,279). Deaths assigned GOS-E=1. GOS-E 2-3 classified as unfavorable neurologic status. GOS-E 4-8 classified as favorable neurologic status. Shaded cells indicate transitions between favorable and unfavorable neurologic status or death.

|  |  | GOS-E Classification  GOS-E at 6-Month Follow-Up | | | | | | | | |  |
| --- | --- | --- | --- | --- | --- | --- | --- | --- | --- | --- | --- |
|  |  | **Favorable**  n=561 (43.9%) | | | | | Unfavorable  n=177 (13.8%) | | Dead n=346 (27.1%) | Missing n=195 (15.2%) |  |
| GOS-E Classification | GOS-E at Hospital Discharge | 8 | 7 | 6 | 5 | 4 | 3 | 2 | 1 | Missing | TOTAL |
| Favorable n=330 (25.8%) | 8 | 57 | 9 | 6 | 3 | 2 | 0 | 0 | 0 | 39 | **116** |
|  | 7 | 17 | 4 | 1 | 3 | 0 | 0 | 0 | 0 | 9 | **34** |
|  | 6 | 20 | 6 | 3 | 6 | 1 | 0 | 0 | 1 | 12 | **49** |
|  | 5 | 4 | 7 | 2 | 1 | 1 | 1 | 0 | 1 | 4 | **21** |
|  | 4 | 35 | 16 | 9 | 11 | 17 | 8 | 0 | 0 | 14 | **110** |
| **Unfavorable**  n=554 (43.3%) | 3 | 78 | 48 | 58 | 44 | 77 | 131 | 1 | 9 | 52 | **498** |
|  | 2 | 1 | 0 | 0 | 0 | 3 | 25 | 6 | 12 | 9 | **56** |
| Dead n=321 (25.1%) | 1 | 0 | 0 | 0 | 0 | 0 | 0 | 0 | 321 | 0 | **321** |
| Missing n=74 (5.8%) | Missing | 6 | 2 | 1 | 1 | 1 | 4 | 1 | 2 | 56 | **74** |
|  | TOTAL | **218** | **92** | **80** | **69** | **102** | **169** | **8** | **346** | **195** | **1,279** |

|  | No change between favorable/unfavorable status or death |
| --- | --- |
|  | Improvement in neurologic status |
|  | Decline in neurologic status or death |

### APPENDIX 6

Sensitivity analysis. Trajectories of neurologic status between hospital discharge and 6-month follow-up among patients enrolled in the Resuscitation Outcomes Consortium Hypertonic Saline Traumatic Brain Injury (TBI) trial. Includes 1,066 of 1,279 TBI subjects with complete neurologic status at both hospital discharge and 6-month follow-up. Neurologic status defined as: favorable GOS-E 4-8, unfavorable GOS-E 2-3, dead GOS-E 1.

| **Neurologic Trajectory**  **(Hospital Discharge 🡪 6-Month Follow-Up)** | **Full Cohort**  **N (%)** | **Hospital Survivors Only**  **N (%)** |
| --- | --- | --- |
| 1) Favorable (GOS-E 4-8) 🡪 Favorable (GOS-E 4-8) | 241 (22.6%) | 241 (32.3%) |
| 2) Favorable (GOS-E 4-8) 🡪 Unfavorable (GOS-E 2-3) | 9 (0.8%) | 9 (1.2%) |
| 3) Favorable (GOS-E 4-8) 🡪 Dead (GOS-E 1) | 2 (0.2%) | 2 (0.3%) |
| 4) Unfavorable (GOS-E 2-3)🡪 Favorable (GOS-E 4-8) | 309 (29.0%) | 309 (41.5%) |
| 5) Unfavorable (GOS-E 2-3)🡪 Unfavorable (GOS-E 2-3) | 163 (15.3%) | 163 (21.9%) |
| 6) Unfavorable (GOS-E 2-3)🡪 Dead (GOS-E 1) | 21 (2.0%) | 21 (2.8%) |
| 7) Dead at Hospital Discharge (GOS-E 1) | 321 (30.1%) | N/A |
